# Association of Digestive Symptoms and Hospitalization in Patients with SARS-CoV-2 Infection

**DOI:** 10.1101/2020.04.23.20076935

**Authors:** George Cholankeril, Alexander Podboy, Vasiliki Irene Aivaliotis, Edward A. Pham, Sean Spencer, Donghee Kim, Ann Hsing, Aijaz Ahmed

## Abstract

**Background:** High rates of concurrent gastrointestinal manifestations have been noted in patients with COVID-19, however the association between these digestive manifestations and need for hospitalization has not been established.

**Methods:** Following expedited approval from our Institutional Review Board, we analyzed retrospectively collected data from consecutive patients with confirmed COVID-19 based on a positive polymerase chain reaction testing at our institution from March 03, 2020 to April 7, 2020. Baseline demographic, clinical, laboratory and patient-reported symptom data were collected at presentation in the emergency room. Multivariable logistic regression analyses were performed to evaluate the association between hospitalization and presence of gastrointestinal symptoms.

**Results:** During this study period, we identified 207 consecutive patients with confirmed COVID-19. 34.5% noted concurrent gastrointestinal symptoms; of which 90% of gastrointestinal symptoms were mild. In a multivariate regression model controlled for demographics and disease severity, an increased risk for hospitalization was noted in patients with any gastrointestinal symptom (adjusted OR 4.84 95% CI: 1.68-13.94]. Diarrhea was associated with a seven-fold higher likelihood for hospitalization (adjusted OR=7.58, 95% CI: 2.49-20.02, P <0.001) and nausea or vomiting had a four times higher odds. (adjusted OR 4.39, 95% CI: 1.61-11.4, P = 0.005).

**Conclusion:** We demonstrate that a significant portion of COVID19 patients have concurrent mild gastrointestinal symptoms and that the presence of these digestive symptoms is associated with a need for hospitalization. With the current focus on streamlining triaging efforts, first responders and frontline providers should consider assessing for digestive symptoms in their initial clinical evaluation and decision-making.

## Background

The current pandemic caused by the severe acute respiratory syndrome coronavirus (SARS-CoV-2), continues to spread globally, and as of April 13, 2020, over 1.7 million cases have been reported worldwide^1^. While respiratory manifestations preponderate in patients with SARS-CoV-2 infection ^2,3^, emerging data suggest a significant prevalence of concurrent gastrointestinal symptomology^4^. Our aim was to examine the association between clinical and disease characteristics, including concurrent digestive manifestations, and need for hospitalization in patients with confirmed COVID-19.

## Methods

Following expedited approval from our Institutional Review Board, we analyzed retrospectively collected data from consecutive patients with confirmed COVID-19 based on a positive polymerase chain reaction testing at our institution from March 03, 2020 to April 7, 2020. Baseline demographic, clinical, laboratory and patient-reported symptom data were collected at presentation. Multivariable logistic regression analyses were performed to assess likelihood for hospitalization with digestive symptoms (nausea/vomiting, diarrhea, abdominal pain, loss of appetite) after adjusting for clinical demographics (age, gender, race/ethnicity), chronic co-morbidities, duration of symptoms, oxygen status, and respiratory symptoms at presentation. Patients with missing covariate data were excluded from the regression model.

## Results

Clinical demographics and characteristics of 207 patients with confirmed COVID-19 are listed in Table 1. Of these 207 patients, 60 patients (29.0%) were hospitalized; with 17 patients (8.2%) requiring intensive care unit (ICU) level of care. To date, there have been four COVID-19 related deaths. Overall, a higher prevalence of males, hypertension, and diabetes mellitus were seen in patients who were hospitalized (P<0.05). Respiratory viral co-infection was found in 14 of 146 (9.1%) tested patients; of whom two patients were hospitalized and three patients had digestive symptoms. Concurrent digestive symptoms were noted in over one-third of all patients, with a higher prevalence observed in those hospitalized to the medical floor and ICU compared those seen only in the emergency room (Table 2). 90% of all digestive symptoms were characterized as mild in severity. Prevalence of acute renal insufficiency was observed to be higher in patients with digestive symptoms than those without digestive symptoms (9.3% versus 3.1%).

After adjusting for confounders and clinical covariates, patients experiencing any digestive symptom (adjusted OR 4.84 95% CI: 1.68-13.94, P < 0.001] had over four-fold higher odds for hospitalization. Diarrhea was associated with a seven-fold higher likelihood for hospitalization (adjusted OR=7.58, 95% CI: 2.49-20.02, P <0.001) and nausea or vomiting had a four times higher odds. (adjusted OR 4.39, 95% CI: 1.61-11.4, P = 0.005).

## Discussion

We demonstrate that a significant portion of COVID19 patients have concurrent mild gastrointestinal symptoms and that the presence of these digestive symptoms is associated with a need for hospitalization. The pathogenesis for gastrointestinal involvement related to SARS-CoV-2 is unknown. However, a critical cellular receptor in the SAR-CoV-2 lifecycle, angiotensin-converting enzyme 2, is abundantly expressed throughout the gastrointestinal tract^5^ and may play a role in worsening digestive symptoms as COVID19 progresses.^6^ Whether digestive symptoms are a surrogate clinical marker for higher levels of viremia or from an alternative pathophysiologic process remains unknown.

There are several limitations to our findings. As this is a retrospective single institution study, our findings may not be broadly generalizable. Also, as this series represents our initial experience treating COVID-19, it is unclear if these results should be viewed on a continuum with changing demographic and clinical information with time. Additionally, due to the short study duration, we were unable to further assess hospitalization outcomes.

In conclusion, while analyzing our initial clinical and demographic data in patients with COVID-19 we identified the presence of gastrointestinal symptoms as a risk factor for higher severity of overall illness and need for hospitalization. With the current focus on streamlining triaging efforts, first responders and frontline providers should consider assessing for digestive symptoms in their initial clinical evaluation and decision-making. Larger prospective studies are needed to validate these observations.

## Data Availability

The datasets generated analysed during the current study are not publicly available due to IRB restraints but are available from the corresponding author on reasonable request.

---

**Appendix Table 1.** Clinical demographics and characteristics of patients with confirmed SARs-CoV-2 infection at Stanford Hospital and Clinics, median [IQR] or No. (%).

|  | All Patients<br>N=207 | Not<br>Hospitalized<br>N=147 | Hospitalized<br>N=60 | Level of Hospitalization |  |
| --- | --- | --- | --- | --- | --- |
|  |  |  |  | Medical Floor<br>N=43 | Intensive Care Unit<br>N=17 |
| **Age** | 49 [34-65] | 43 [ 31 – 58]* | 62 [43 – 77] | 65 [45-77] | 55 [42 – 70] |
| Male | 104 (50.2) | 72 (49.0)* | 32 (53.3) | 22(51.2) | 10 (58.8) |
| Female | 103( 49.8) | 75 (51.0)* | 28 (46.7) | 21(48.8) | 7(41.2) |
| **Race/Ethnicity** |  |  |  |  |  |
| Caucasian | 87 (42.4) | 66(44.9) | 21 (36.2) | 18(41.9) | 3 (20.0) |
| Asian | 42 (20.5) | 27 (18.4) | 15 (25.9) | 11 (25.6) | 4 (26.7) |
| Hispanic | 62 (30.2) | 45 (30.6) | 17 (29.3) | 9 (20.9) | 8 (53.3) |
| Black | 2 (1.0) | 1 (0.7) | 1 (1.7) | 1 (2.3) | 0 (0.0) |
| Other | 12 (5.9) | 8 (5.4) | 4 (6.9) | 4 (9.3) | 0 (0.0) |
| **Body Mass Index (kg/m<sup>2</sup>)** | 26 .3 [23.2-31.0] | 25.4 [23.1 – 30.8] | 28.0 [24.0 – 32.5] | 27.0 [23.8-31.2] | 29.5 [27.5 – 33.8] |
| **Current Smoker** | 3 (1.6) | 1 (0.7) | 2 (3.5) | 2 (4.8) | 0 (0.0) |
| **History of Recent Travel** |  |  |  |  |  |
| Domestic | 20 (10.1) | 16 (11.4) | 4 (6.8) | 1 (2.3) | 3 (18.8) |
| International | 20 (10.1) | 16 (11.4) | 4 (6.8) | 4 (9.3) | 0 (0.0) |
| Cruise | 6 (3.0) | 4 (2.9) | 2 (3.4) | 1 (2.3) | 1(6.3) |
| **Healthcare worker** | 22(10.6)) | 16 (10.8) | 6 (10.2) | 3 (7.0) | 3 (17.6) |
| **Known Exposure to COVID** | 73 (36.8) | 52 (37.1) | 21 (36.2) | 14 (33.3) | 7 (43.8) |
| **Past Medical History** |  |  |  |  |  |
| Chronic Liver Disease | 5 (2.7) | 4 (3.1) | 1 (1.7) | 1 (2.3) | 0 (0.0) |
| Chronic Pulmonary Disorder | 42 (20.3) | 25 (17.0) | 17 (28.3) | 13 (30.2) | 4 (23.5) |
| Hypertension | 52 (25.5) | 30 (20.7)* | 22 (37.3) | 14 (33.3) | 8 (47.1) |
| Diabetes | 33 (16.0) | 16 (11.0)* | 17 (28.3) | 10 (23.3) | 7 (41.2) |
| Cardiovascular Disease | 24 (11.7) | 13 (8.9) | 11 (18.3) | 10(23.3) | 1 (5.9) |
| Metabolic Syndrome | 19 (9.2) | 8 (5.5)* | 11 (18.3) | 8 (18.6) | 3 (17.7) |
| Chronic Kidney Disease | 10 (4.4) | 2 (1.4) | 8 (13.3) | 7 (16.3) | 1 (5.9) |
| **Medication Use** |  |  |  |  |  |
| angiotensin converting enzyme inhibitors/ angiotensin-receptor blockers | 23 (11.2) | 14 (9.6) | 9 (15.0) | 5 (11.6) | 4 (23.5) |
| Chronic immunosuppression | 7 (3.4) | 4 (2.7) | 3 (5.0) | 3 (7.0) | 0 (0.0) |
| Immunotherapy | 5 (2.4) | 3 (2.1) | 2 (3.33) | 1 (2.3) | 1 (5.9) |
| **Respiratory or Viral Symptoms** |  |  |  |  |  |
| Fever | 142 (68.6) | 92 (62.6)* | 50 (83.3) | 36 (83.7) | 14 (82.4) |
| Cough | 175 (85.4) | 118 (80.8)* | 57 (96.6) | 41 (95.6) | 16 (94.1) |
| Shortness of Breath | 108 (52.2) | 58 (39.5)* | 50 (83.3) | 34 (79.0) | 16 (94.1) |
| Sore Throat | 54 (26.2) | 43 (29.3) | 11 (18.6) | 6 (14.3) | 5 (29.4) |
| Myalgias | 105 (51.0) | 68 (46.6) | 37 (61.7) | 23 (53.5) | 14 (82.4) |
| Duration of respiratory viral symptoms, days | 5 [3-7] | 5 [3-7]* | 7 [3.5 – 9] | 7 [3-9] | 7 [6-8] |
| **Gastrointestinal Symptoms** |  |  |  |  |  |
| Any Gastrointestinal Symptom<sup>a</sup> | 70 (34.5) | 34 (23.5)* | 28 (46.7) | 26 (63.1) | 10 (58.2) |
| Nausea or Vomiting only | 22 (10.8) | 14 (9.6)* | 8 (13.8) | 6 (14.6) | 2 (11.8) |
| Diarrhea only | 22 (10.8) | 10 (6.9)* | 12 (20.7) | 8 (19.5) | 4 (23.5) |
| Nausea or vomiting and diarrhea | 10 (4.9) | 3 (2.1) | 7 (12.1) | 5 (12.2) | 2 (11.8) |
| Abdominal Pain | 14 (7.1) | 10 (7.1) | 4 (7.0) | 4 (10.0) | 0 |
| Duration of Gastrointestinal symptoms, days | 1 [0-4] | 1 [0-3] | 2 [1- 4] | 2 [1-4] | 4 [2-7] |

| Laboratory values n=115 |  |  |  |  |  |
| --- | --- | --- | --- | --- | --- |
| White blood cell count (K/uL) | 5.6 [4.1-7.3] | 5.8 [4.5-7.2] | 5.4 [3.9 – 7.8] | 5.2 [3.8-8.2] | 5.3 [3.9-7.0] |
| Absolute lymphocyte count (K/uL) | 0.9 [0.7-1.5] | 1.1 [0.8-1.7] | 0.9 [ 0.6 – 1.2] | 0.9 [0.6-1.2] | 0.6 [0.4-1.0] |
| Platelet count (K/uL) | 190 [159-241] | 203 [169-244] | 182 [153 – 241] | 183 [142-241] | 181 [157-250] |
| Serum Sodium (mmol/L) | 138 [135-141] | 139 [136-141]* | 136 [133.5 – 139] | 136 [132-139] | 136 [134-138] |
| Serum Creatinine (mg/dL) | 0.8 [0.6-1.0] | 0.8 [0.6-0.9]* | 0.9 [0.7 -1.1] | 0.9 [0.8-1.1] | 0.8 [ 0.5-1.0] |

\* $P < 0.05$ not hospitalized vs hospitalized.
<sup>a</sup> Gastrointestinal symptoms include nausea or vomiting, diarrhea, abdominal pain, or loss of appetite

**Table 2.**
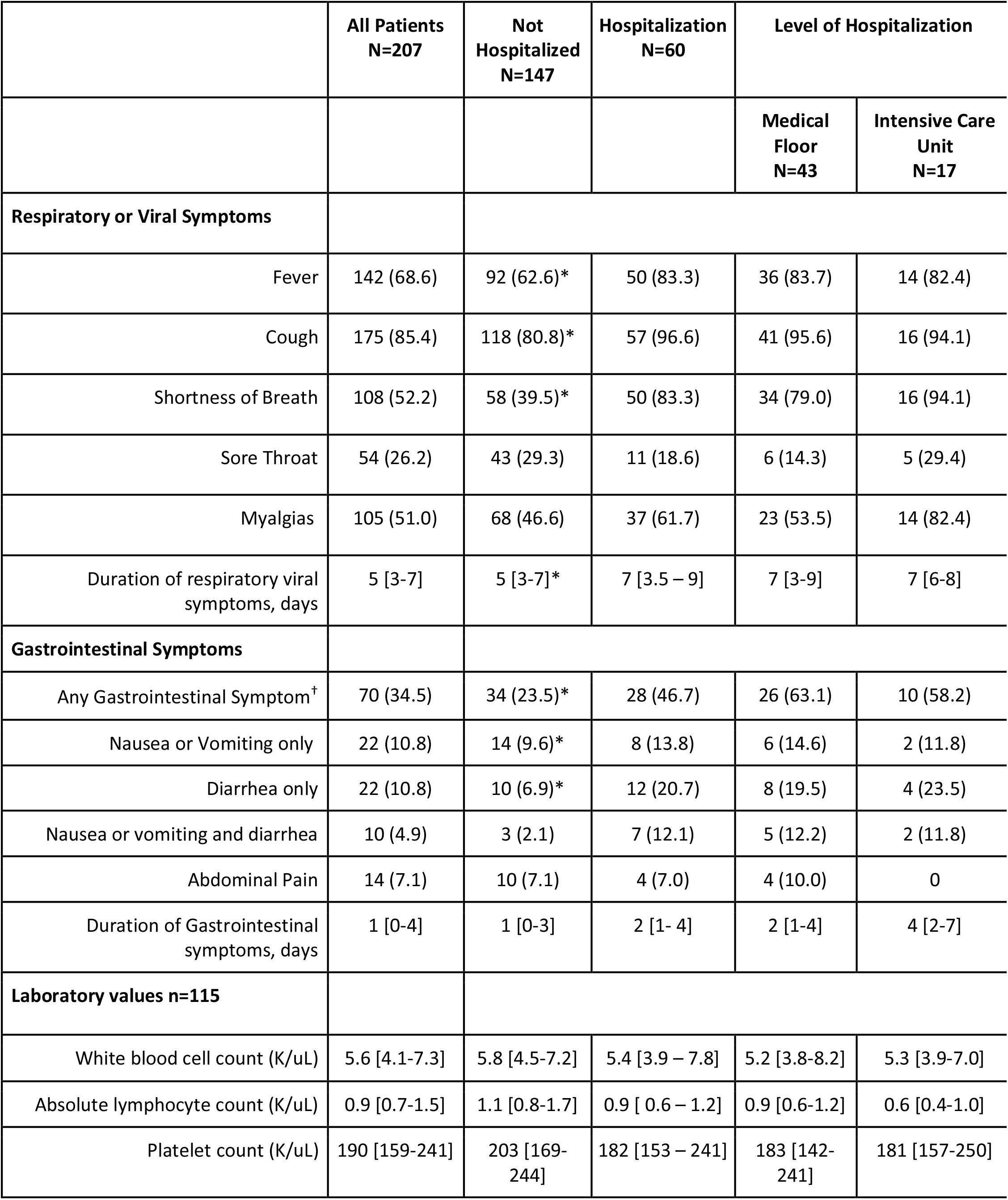
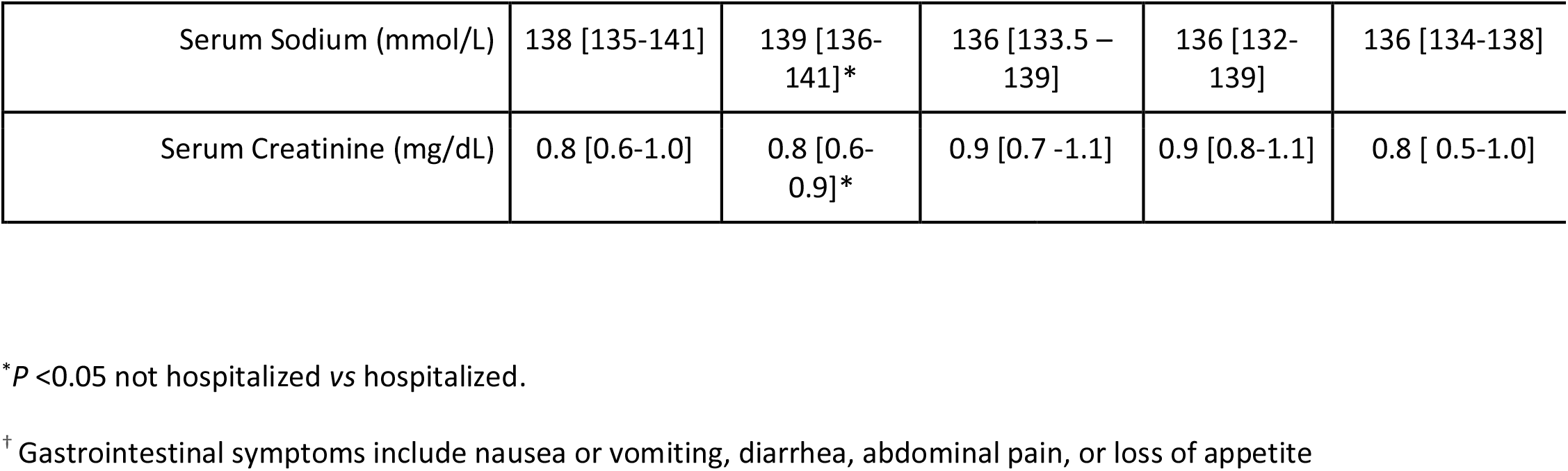
Clinical presentation of respiratory and gastrointestinal symptoms and laboratory findings at initial evaluation in patients with confirmed SARS-CoV-2 Infection, median [IQR] or No. (%).

## Author Contributions

A.P. and G.C. equally contributed to this paper with conception and design of the study, literature review and analysis, drafting and critical revision and editing, and final approval of the final version. V.A., A.P., E.A.P., S.P., and B.T. assisted in data acquisition, manuscript preparation and critical appraisal of the manuscript. A.A. and D.K. provided critical appraisal of the manuscript.

